# Transforming Patient Voices into Early Predictors of Survival Using Nonlinear Mixed-Effect Models and AI/ML for Patient-Centered Decision-Making

**DOI:** 10.64898/2026.04.30.26352154

**Authors:** Congyu Zhang, Peng Xia, Wanbing Wang, Ghassan Slim, Benyam Muluneh, Jennifer Jansen, Lynne I. Wagner, William A. Wood, Huaxiu Yao, Jim H. Hughes, Ethan Basch, Jiawei Zhou

## Abstract

Patient-reported outcomes (PROs) capture the patient voice and have been associated with improved clinical outcomes in oncology, but their prognostic and predictive value remains underutilized due to challenges in interpreting these highly variable and noisy PRO data. Here, we developed a quantitative modeling framework integrating nonlinear mixed-effects (NLME) and item response theory (IRT) to characterize symptom-level PRO trajectories and transform them into clinically actionable predictors. Using longitudinal PRO data from 589 patients with metastatic cancers in the PRO-TECT trial, we modeled 332,920 symptom responses to estimate patient-specific PRO trajectory parameters while accounting for variability and noise. IRT-NLME modeling captured heterogeneous symptom-level PRO dynamics and is more informative than modeling with composite PRO scores. PRO trajectory parameters were strongly associated with overall survival, acute care utilization, and treatment modifications. Machine learning models leveraging these parameters achieved robust prediction of survival (AUC-ROC 0.80) and retained prognostic performance using the first 30 – 180 days of PRO observations, with AUCs of 0.69-0.78. Similar predictive performance was observed for hospitalization (AUC 0.75), emergency department visit (AUC 0.65), treatment discontinuation (AUC 0.71), and dose reduction (AUC 0.67). These findings demonstrate that longitudinal PRO trajectories can serve as early, patient-centered biomarkers of clinical risk. By converting complex symptom data into interpretable and predictive metrics, this quantitative framework provides a practical pathway to integrate the patient voice into clinical decision-making and advance precision oncology.

ClinicalTrials.gov registration: NCT03249090

## Introduction

Listening to the patient’s voice is fundamental to delivering high-quality care, particularly in oncology, where treatments often carry substantial toxicity and impact on quality of life^1^.

Patient-reported outcomes (PROs) provide a direct and systematic way to capture the patient voice, reflecting patients’ experiences of symptoms, treatment toxicity, and functional well-being—dimensions that are often under-recognized in routine clinical assessments^2^.

Prior studies have shown that routine PRO monitoring can improve symptom control, reduce emergency department (ED) visits and hospitalizations^3,4^, and even improve survival^5^. In addition, evidence suggests that PRO measurements have prognostic value for survival in patients with cancer^6^. These findings demonstrate that PRO monitoring is closely associated with improved clinical outcomes and can directly inform patient care.

Despite their clinical value, the use of PROs in routine practice and decision-making remains limited. A major challenge lies in the complexity of longitudinal PRO data, which are often characterized by substantial variability and noise^7–9^. These features make it difficult to distinguish clinically meaningful symptom changes from random fluctuations and to reliably interpret PRO trends over time. Furthermore, PRO questionnaires usually measure multiple different symptoms, but it is not clear how to interpret results when some symptoms improve or worsen while others remain stable, or whether they should be combined into a composite score^10^. In addition, missing data are common in PRO collection, as patients may skip individual items or be unable to complete assessments due to clinical deterioration^11,12^. Existing approaches often rely on imputing missing values, which can introduce bias and reduce confidence in the results^13^. Together, these challenges have hindered the interpretation and prognostic ability of PRO data, limiting their broader implementation in oncology care.

In this study, we have developed a quantitative modeling framework integrating nonlinear mixed-effects (NLME)^14^ and item response theory (IRT)^15^ to address these challenges and characterize longitudinal, symptom-level PRO trajectories. This approach extracts clinically meaningful patterns, distinguishing them from variability and noise. Notably, this approach jointly assesses all symptoms while preserving symptom-specific changes over time.

Additionally, it handles missing data without explicit imputation assumptions, as reported in previous studies^16,17^. We applied this framework to derive patient-specific PRO trajectory parameters and evaluated their ability to predict key clinical outcomes, including overall survival (OS), acute care utilization, and treatment modifications. Importantly, we tested whether PRO trajectories estimated using early observation data can predict subsequent clinical outcomes, demonstrating the potential of PROs as early indicators for risk stratification and timely clinical intervention. Together, this work establishes a pathway to transform patient-reported data into clinically actionable, patient-centered biomarkers that can support precision oncology.

## Results

### Study Data Description

Weekly PRO data comprising 332,920 PRO responses were collected from 589 patients with metastatic cancers enrolled in the PRO-TECT trial (NCT03249090)^4^. Patient demographics and clinical characteristics are summarized in **Table 1**. Clinical outcomes were collected, including mortality, acute care events (hospitalization and ED visits), and treatment modifications (treatment discontinuation and dose reduction). Median time-to-event outcomes are summarized in **Supplementary Table 1**. Multivariable logistic regression models were used to evaluate associations between baseline demographics, clinical characteristics, and clinical outcomes (**Fig. 1a** and **Supplementary Fig. 1**). Notably, ECOG performance status was the only factor significantly associated with OS (hazard ratio, 1.82; 95% CI, 1.29–2.67; P < 0.001). Kaplan-Meier curves for OS are shown in **Fig. 1b**.

**Figure 1.**
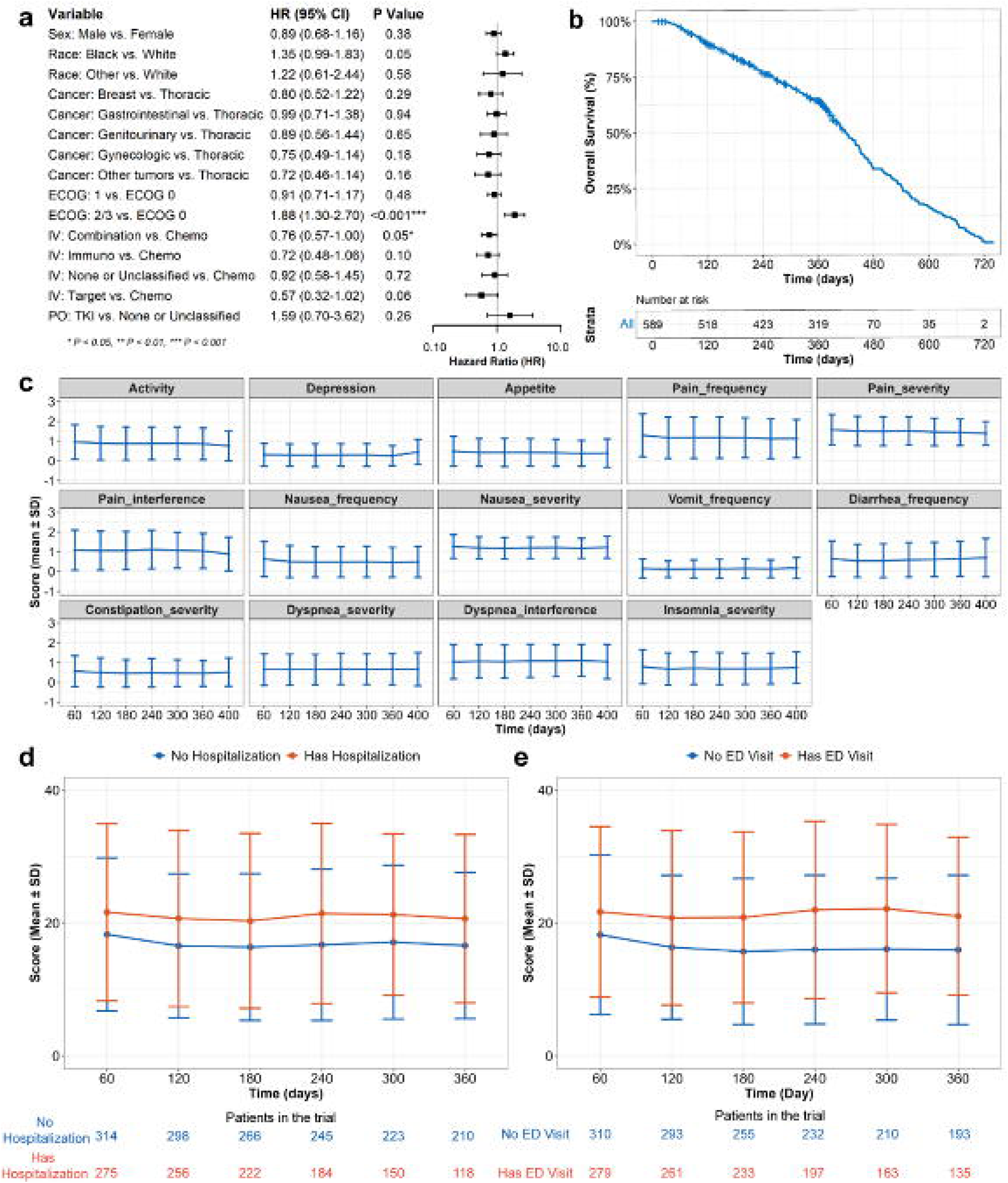
Baseline characteristics and longitudinal patient-reported outcome (PRO) scores in the PRO-TECT trial. **(a)** Associations between baseline clinical characteristics and overall survival estimated using Cox proportional hazards models. **(b)** Kaplan-Meier curve for overall survival of patients in the study cohort. The number of patients at risk is shown below. **(c)** Longitudinal PRO symptom scores (mean ± s.d.) over time in the PRO-TECT trial. Scores were summarized within each 60-day time window, except for the final interval, which was defined as 40 days. **(d)** Longitudinal PRO total scores (mean ± s.d.) in patients with and without hospitalization events. **(e)** Longitudinal PRO total scores (mean ± s.d.) in patients with and without ED visits. ECOG, Eastern Cooperative Oncology Group; s.d., standard deviation; ED, emergency department; IV, intravenous treatment; PO, oral treatment; TKI, tyrosine kinase inhibitors; PRO, Patient-Reported Outcomes.

**Table 1.** The demographics information of the patients in the analysis.

| **Dataset** | Training (N=471) | Testing (N=118) | Total (N = 589) |
| --- | --- | --- | --- |
| **Age, median(range), years** | 64 (29, 89) | 64 (30, 88) | 64 (29, 89) |
| **Sex** |  |  |  |
| Male | 171 (36.3%) | 61 (51.7%) | 232 (39.4%) |
| Female | 300 (63.7%) | 57 (48.3%) | 357 (60.6%) |
| **Race** |  |  |  |
| Native American | 8 (1.7%) | 3 (2.5%) | 11 (1.9%) |
| Asian | 2 (0.4%) | 0 | 2 (0.3%) |
| Black | 83 (17.8%) | 15 (12.7%) | 98 (16.8%) |
| Native Hawaiian | 2 (0.4%) | 0 | 2 (0.3%) |
| White | 370 (79.4%) | 100 (84.7%) | 470 (80.5%) |
| Multiple Races Selected | 1 (0.2%) | 0 | 1 (0.2%) |
| Missing | 5 (1.1%) | 0 | 5 (0.8%) |
| **Ethnicity (Hispanic or Latino)** | 9 (1.9%) | 5 (4.2%) | 14 (2.4%) |
| **Cancer Type** |  |  |  |
| Thoracic (Lung, Thymus) | 96 (20.4%) | 22 (18.6%) | 118 (20%) |
| Breast | 83 (17.6%) | 14 (11.9%) | 97 (16.5%) |
| Colorectal, Anal | 75 (15.9%) | 24 (20.3%) | 99 (16.8%) |
| Prostate | 22 (4.7%) | 9 (7.6%) | 31 (5.3%) |
| Gynecologic (Ovarian, Cervix, | 50 (10.6%) | 13 (11.0%) | 63 (10.7%) |
| Myeloma, Lymphoma | 26 (5.5%) | 5 (4.2%) | 31 (5.3%) |
| Melanoma, Skin | 10 (2.1%) | 1 (0.8%) | 11 (1.9%) |
| Genitourinary Non-prostate | 29 (6.2%) | 7 (5.9%) | 36 (6.1%) |
| Gastro-esophageal, small bowel | 20 (4.2%) | 36 (6.1%) | 25 (4.2%) |
| Pancreas, Hepatobiliary | 34 (7.2%) | 36 (6.1%) | 48 (8.1%) |
| Other (Brain, Head/Neck, Thyroid,<br>Sarcoma, Other soft tissue,<br>Unknown primary) | 26 (5.5%) | 4 (3.4%) | 30 (5.1%) |
| **ECOG** |  |  |  |
| 0 | 203 (43.1%) | 48 (40.7%) | 251(42.6%) |
| 1 | 217 (46.1%) | 58 (49.2%) | 275 (46.7%) |
| 2 | 46 (9.8%) | 12 (10.2%) | 58 (9.8%) |
| 3 | 5 (1.1%) | 0 | 5 (0.8%) |
| **Intravenous Treatment** |  |  |  |
| Chemotherapy Only | 179 (38%) | 49 (41.5%) | 228 (38.7%) |
| Targeted therapy Only | 36 (7.6%) | 9 (7.6%) | 45 (7.6%) |
| Immunotherapy Only | 75 (15.9%) | 17 (14.4%) | 92 (15.6%) |
| Combination Therapy | 110 (23.4%) | 31 (26.3%) | 141 (23.9%) |
| None or Unclassified | 71 (15.1%) | 12 (10.2%) | 83 (14.1%) |
| **Oral Treatment** |  |  |  |
| Tyrosine Kinases Inhibitors | 23 (4.9%) | 2 (1.7%) | 25 (4.2%) |
| None or Unclassified | 448 (95.1%) | 116 (98.3%) | 564 (95.8%) |

The PRO dataset included 14 symptom items assessing treatment-related toxicity and disease burden (**Supplementary Table 2**). The overall missingness rate was 23.8%, with no significant change over time. (**Supplementary Fig. 2**). PRO scores over time for individual symptoms are shown in **Fig. 1c**. Longitudinal total PRO scores were further examined after stratification by clinical outcomes, including hospitalization (**Fig. 1d**), ED visits (**Fig. 1e**), and treatment modification (**Supplementary Fig. 3**).

### Nonlinear mixed-effects (NLME) modeling captured longitudinal PRO trajectories

We developed a quantitative framework to characterize longitudinal PRO trajectories and predict clinical outcomes (**Fig. 2a**). A semi-mechanistic NLME model was constructed to describe PRO trajectories during cancer therapy, capturing patients’ symptom changes driven by treatment effects, toxicity, and disease burden (**Fig. 2b**). This NLME modeling approach explicitly accounts for substantial variability and noise in longitudinal PRO data, allowing estimation of patient-specific trajectory parameters, and separating clinically meaningful changes in symptoms from non-clinical related fluctuations (residual error). (**Fig. 2c**).

**Figure 2.**
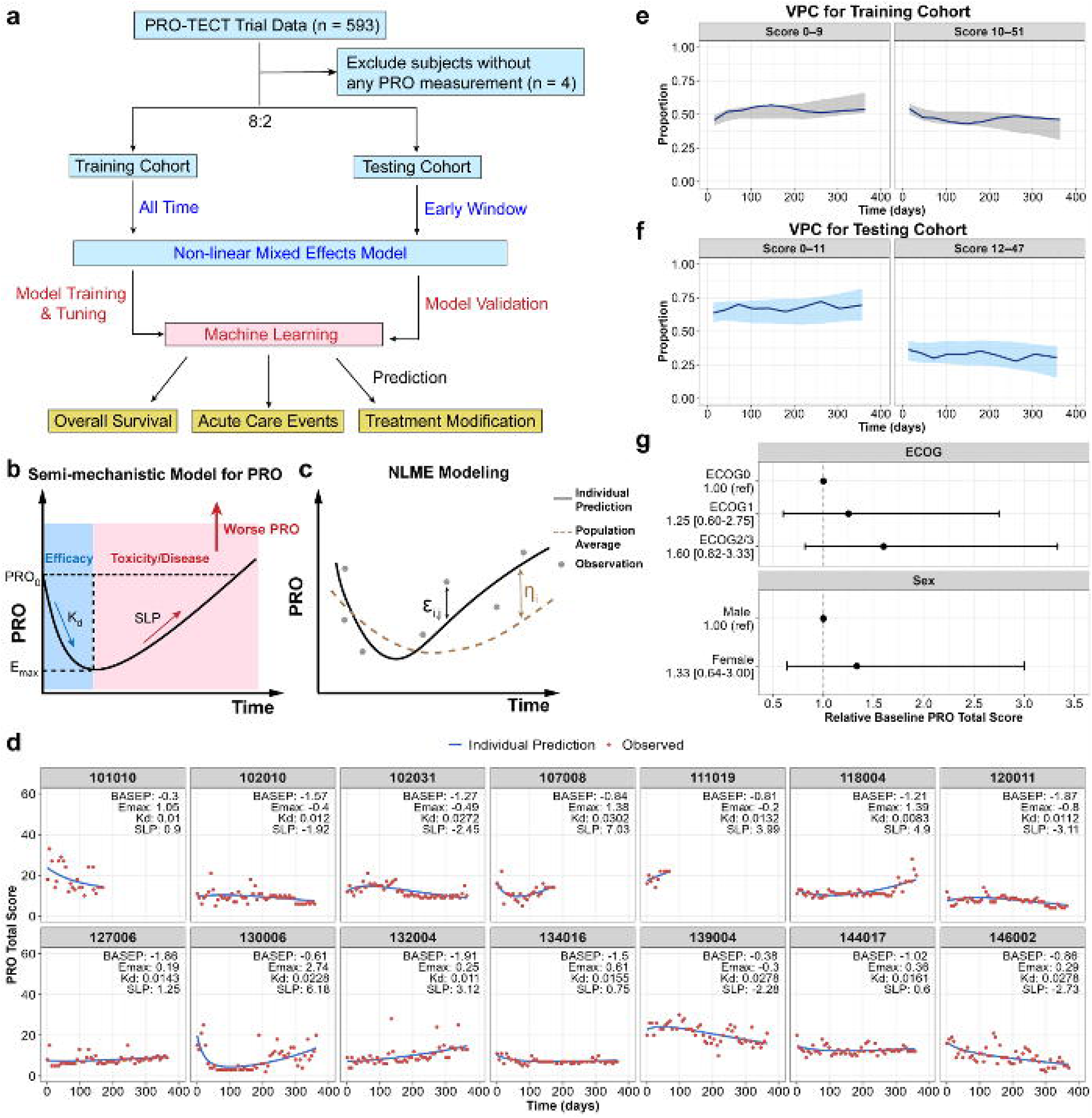
Nonlinear mixed-effects (NLME) model captured longitudinal trajectories of the patient-reported outcomes (PRO) total score at both the population-average and individual-participant levels. (a) Study workflow. (b) Semi-mechanistic model describing longitudinal PRO trajectories during treatment, capturing the combined effects of treatment efficacy, treatment-related toxicity and disease burden on PROs. (c) Diagram of NLME modeling approach. All available longitudinal PRO data from individual participants were analyzed simultaneously in the PRO trajectory model (shown in b). This approach estimates both the average trajectory across the study population and variability in the data. The term 1J_i_ represents how much an individual patient’s PRO trajectory differs from the population average; and E_i,j_, represents random variation (residual error) within the same subject (subject ***i***) at measurement time point ***j***. (d) Representative longitudinal PRO trajectories for 14 randomly selected participants. Red circles represent observed data, and solid blue lines represent model-predicted PRO trajectories. (e) VPC evaluates the training cohort NLME model performance. VPC is a standard diagnostic used to assess how well the model captures the observed data. Solid lines indicate the median observed PRO total scores over time, and shaded areas represent the 95% prediction interval from the model. Good agreement between the shaded regions and the observed data indicates that the model adequately captures the underlying data patterns. (f) VPC evaluates testing cohort NLME model performance. (g) Forest plot showing the effects of ECOG performance status and sex on baseline PRO total score. Circles represent estimated effect sizes, and horizontal bars indicate 95% confidence intervals. Estimated individual PRO trajectory parameters are shown for each patient. VPC, visual predictive check.

To ensure we both develop and validate the model, the dataset was randomly split into training and testing cohorts (80:20). NLME models were trained using all available PRO data in the training cohort and externally validated in the testing cohort. Distributions of clinical outcomes were generally comparable between the training and testing cohorts **(Supplementary Table S1)**, indicating no substantial imbalance introduced by random data splitting.

We first built NLME models using total PRO scores aggregated across all 14 symptoms.

The NLME model captured the observed total PRO scores at both the individual-patient and population-average levels. At the individual-patient level, the model described diverse patient-specific PRO trajectories and estimated distinct trajectory parameters for each patient (**Fig. 2d**). At the population-average level, the model captured the median trajectory patterns across patients in both the training and testing cohorts (**Fig. 2e-f**). ECOG performance status and sex were associated with baseline PRO levels, with higher PRO scores (indicating worse symptom burden) observed in patients with ECOG 2-3 versus ECOG 0 (relative ratio, 1.6; 95% confidence interval [CI], 0.82–3.33), and in female versus male patients (relative ratio, 1.33; 95% CI, 0.64–3.0, **Fig. 2g**). Final model parameter estimates are provided in **Supplementary Table S3**.

### Embedding item-response theory (IRT) into NLME modeling to capture symptom-level PRO trajectories

Although NLME models of total PRO scores captured the overall trajectory, the use of aggregate measures mask symptom-level heterogeneity. To address this, we extended the NLME framework and incorporated an IRT component, enabling joint modeling of individual symptoms through shared underlying disease domains (**Fig. 3a**). Within this framework, symptom responses were linked to an underlying disease severity using symptom-specific parameters, allowing each symptom to reflect disease severity differently with different deterioration rates (**Fig 3b-c**). This approach can capture heterogeneous, symptom-specific PRO trajectories over time while preserving the population-average and individual-level structure of the NLME framework. We applied this framework to estimate PRO trajectories from longitudinal symptom-level PRO data. Model predictions closely matched the observed symptom trajectories, supporting the model’s ability to capture symptom-specific PRO dynamics (**Fig. 3d**). Final parameter estimates for the IRT-NLME model are provided in **Supplementary Table S4-5**.

**Figure 3.**
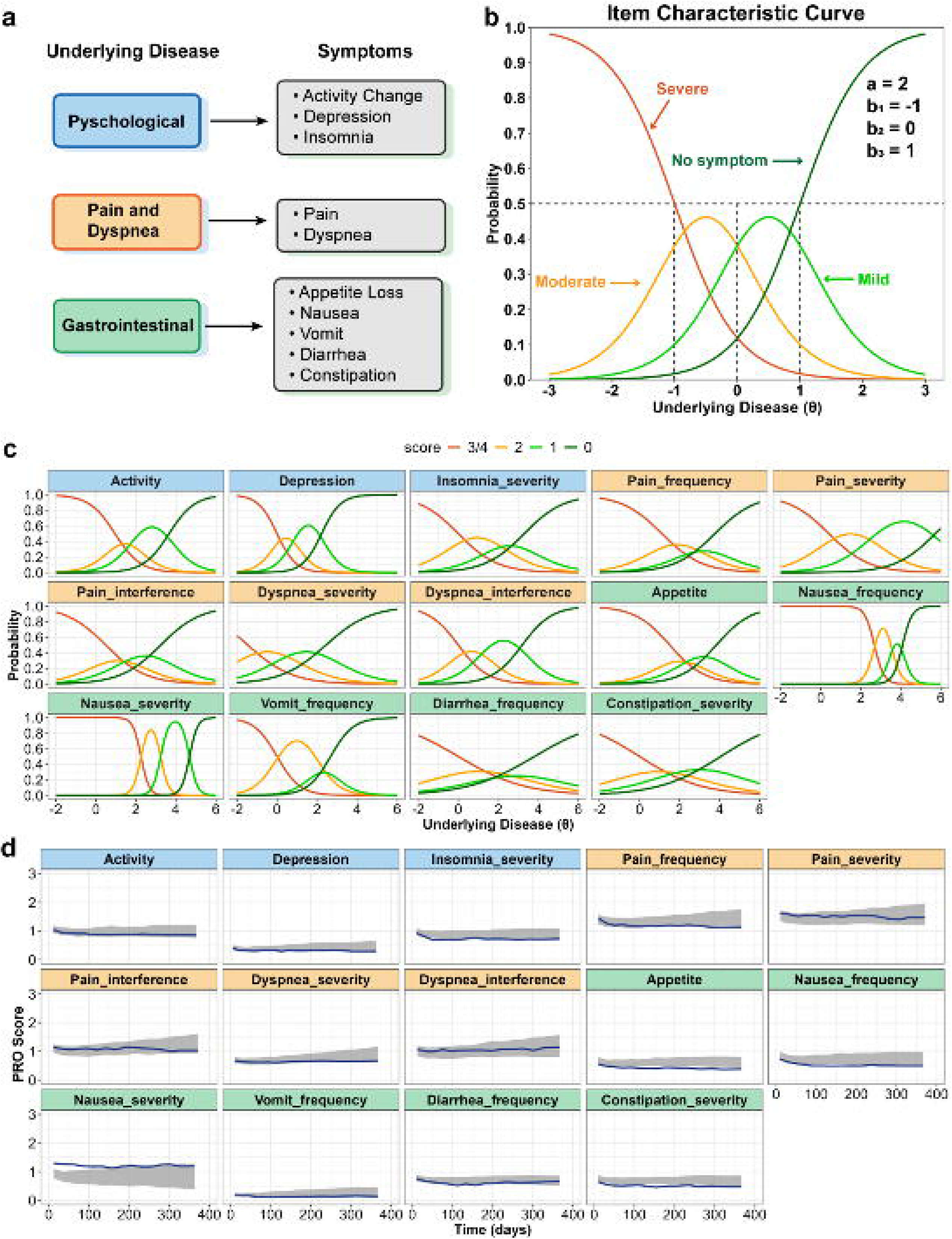
Extending nonlinear mixed-effects (NLME) model with item response theory (IRT) enables symptom-level modeling of patient-reported outcome (PRO) trajectories. (a) Conceptual framework linking individual patient-reported symptoms to underlying disease domains. Symptoms were grouped into three latent domains representing shared underlying disease processes, a common approach in IRT to capture correlated symptom trajectories. These underlying disease domains were subsequently modeled using the NLME approach to capture both population-level trends and individual-level trajectories, as shown in Figure 2. (b) ICC showing the relationships between underlying disease severity and symptom response levels in the IRT framework. In this diagram, the horizontal axis represents the underlying disease severity (θ), and the vertical axis represents the probability of each symptom response level (for example, no symptom, mild, moderate, and severe). The same θ was assigned to all symptoms within each disease domain. Each curve corresponds to one response level, showing how likely a patient is to report that level at a given disease severity. As underlying disease severity increases (θ decrease), the probability shifts from lower to higher response levels, indicating worsening symptoms. This relationship is governed by two symptom-specific parameters: the discrimination parameter (a), which reflects how strongly the symptom is associated with disease severity, and the difficulty parameters (b_1_ – b_3_), which mark when patients transition between adjacent symptom levels. (c) ICC for individual symptoms estimated from the IRT model. (d) VPC evaluates the performance of the NLME model with the IRT extension. Solid lines represent the median observed PRO scores over time for each symptom, and shaded areas indicate the 95% model-predicted intervals. VPC, visual predictive check. ICC, item characteristic curve. Panels’ color corresponds to the symptom domain: blue for psychological, orange for pain/dyspnea, and green for gastrointestinal.

We next validated the model using PRO data from the independent testing cohort. Using all available PRO data, the model accurately recapitulated longitudinal PRO trajectories across all symptoms in the testing cohort (**Supplementary Fig. S4**). To further evaluate model generalizability in early observation data, we assessed model performance using PRO data restricted to the first 30, 60, 90, and 180 days, and compared the resulting trajectories with observed PRO profiles in the testing cohort. The model maintained strong performance across all symptoms, even when using PRO data from early observation windows (**Supplementary Fig. S5**). These findings demonstrate that the IRT-NLME framework can reliably reconstruct PRO trajectories using only early longitudinal data, supporting its validity and potential utility for early prediction of PROs.

### Symptom-level PROs provide more informative signals than total PRO scores

We compared trajectory parameters derived from total PRO score NLME models with those from symptom-level IRT-NLME models. At baseline, the total PRO score did not differ significantly across cancer types (p = 0.15, **Fig. 4a**). In contrast, symptom-level analyses revealed distinct patterns: patients with breast and gastrointestinal (GI) cancers had significantly worse baseline symptom burden in GI domain than those with genitourinary cancers (p = 0.013 and 0.002; **Fig. 4b**). In addition, patients with thoracic cancers exhibited higher baseline symptom burden in pain and dyspnea domains compared to those with GI cancers (p = 0.017; **Fig. 4c**).

**Figure 4:**
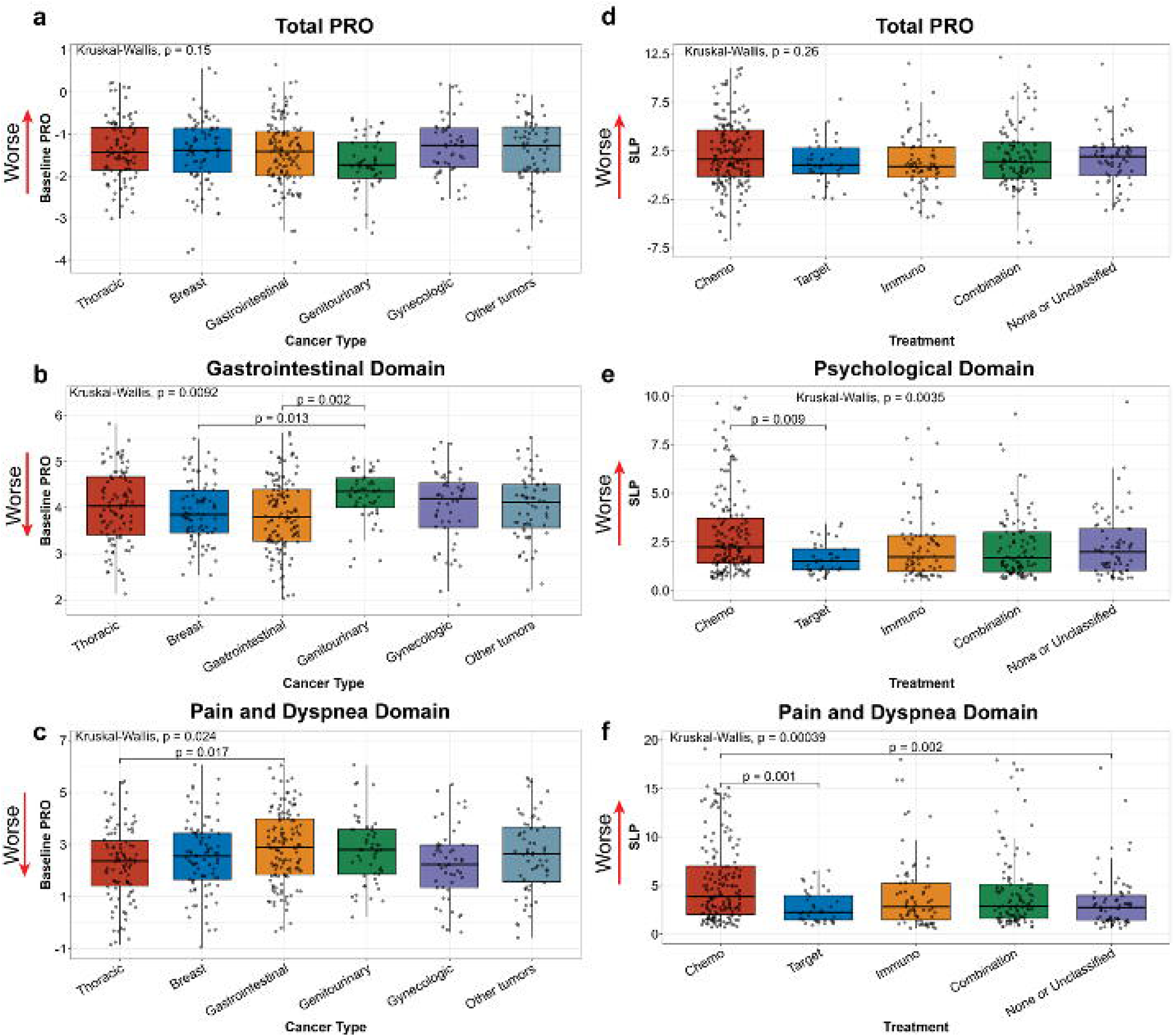
Comparison of patient-reported outcome (PRO) trajectory parameters obtained from total PRO score or symptom-level PRO across cancer types and treatments. (a–c) Comparison of baseline PRO parameter across cancer types for total PRO score model (a), gastrointestinal symptoms domain from IRT model (b), and pain and dyspnea symptoms domain from IRT model(c). (d–f) Comparison of PRO deterioration rate across treatment groups for PRO total score model(d), psychological symptoms domain from IRT model (e), and pain and dyspnea symptoms domain from IRT model(f). Kruskal-Wallis’s test was applied for group comparison and Dunn‘s post hoc test for pairwise comparisons. Red arrows indicate the direction of worse symptoms burden of the patients. IRT, item-response theory.

Similarly, no differences in the median of PRO deterioration rates were observed across treatment types when using total PRO scores (p = 0.26, **Fig. 4d**). However, symptom-level PRO modeling identified significantly faster deterioration rate with chemotherapy compared to targeted therapies, particularly in psychological domains (p = 0.009; **Fig. 4e**) and in pain and dyspnea domains (p = 0.001; **Fig. 4f**). Together, these findings demonstrate that symptom-level PROs provide more informative signals than composite PROs, highlighting the value of the IRT-NLME model for capturing symptom-level PRO trajectories.

### PRO trajectories enable early prediction of overall survival (OS)

We first examined the associations between PRO trajectory parameters derived from the symptom-level IRT-NLME model and key clinical outcomes, including OS, acute care utilization, and treatment modifications. Notably, faster rates of PRO deterioration and higher baseline symptom burdens were significantly associated with increased risk of death, hospitalization, and treatment discontinuation (**Fig. 5a**). Together, these findings demonstrate that longitudinal PRO trajectories capture clinically meaningful signals linked to clinical outcomes, supporting their use as predictors of survival and healthcare utilization.

**Figure 5.**
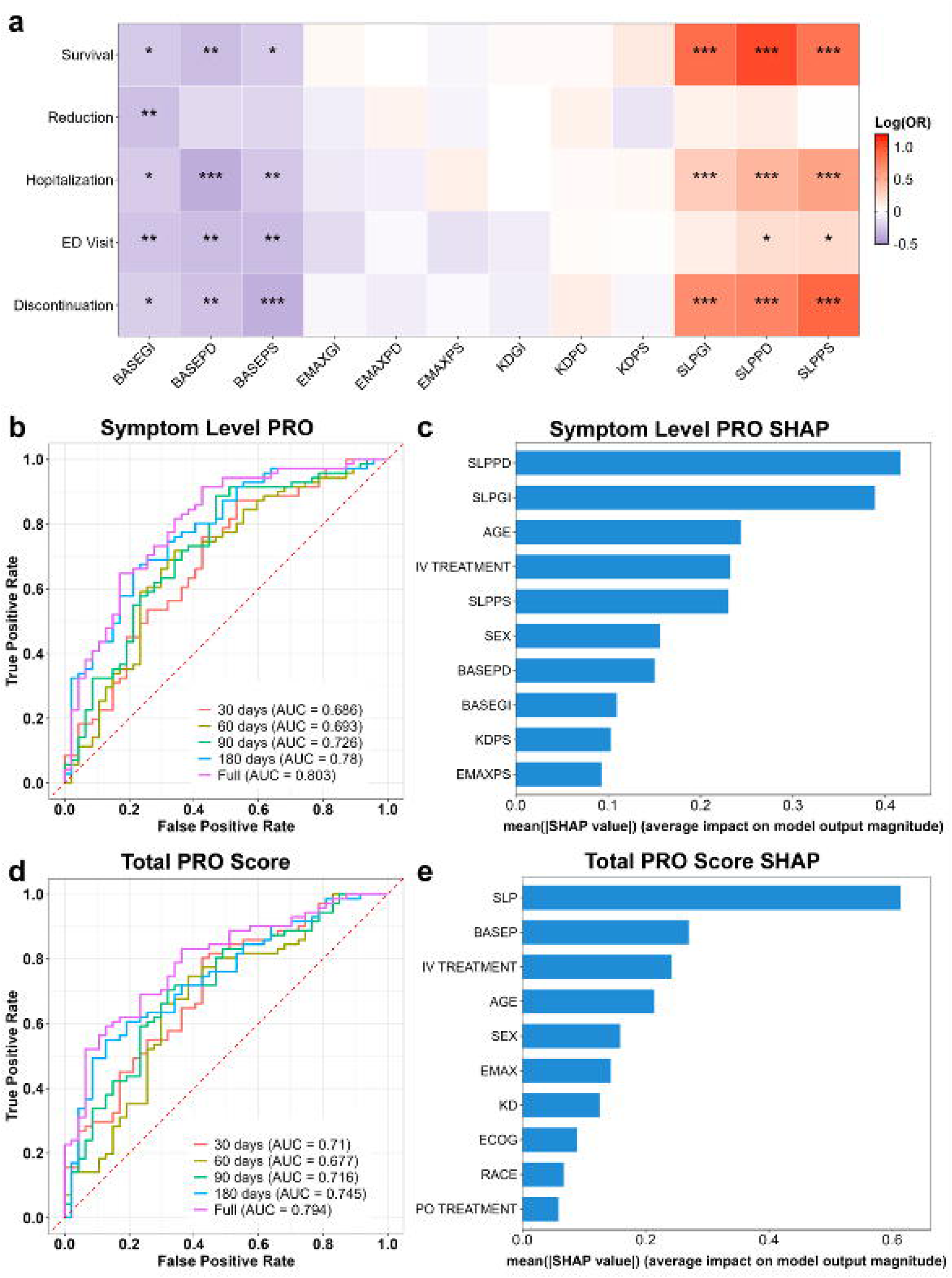
PRO trajectories enable early prediction of overall survival. (a) Heatmap shows the association between PRO trajectory parameters and clinical outcomes based on univariable logistic regression models. (b) ROC curves for machine learning models predicting OS using PRO trajectory parameters derived from the first 30, 60, 90, and 180 days or all available PRO data. (c) Feature importance of the machine learning model in (b), quantified using SHAP values. (d) ROC curves for machine learning models predicting OS using PRO trajectory parameters. (e) Feature importance of the machine learning model in (d), quantified using SHAP values. In (a–c), PRO trajectory parameters were derived from IRT-extended NLME models at the symptom level. In (d–e), PRO trajectory parameters were derived from NLME models based on total PRO scores. GI (PD, PS), gastrointestinal, pain/dyspnea and psychological domains. BASE, baseline symptom severity. EMAX, maximum symptom improvements. KD, symptom improvement rate. SLP, symptom deterioration rate. OR, odds ratio. NLME, nonlinear mixed effect. IRT, item-response theory. ROC, receiver-operating characteristic. OS, overall survival. AUC, area under the curve. SHAP, Shapley Additive Explanations. P<0.05, *; p<0.01, **; p<0.001, ***

PRO trajectory parameters were then used as inputs to machine learning models to predict OS. Models trained on symptom-level PRO trajectories from the training cohort and validated in the testing cohort demonstrated good predictive performance (area under the receiver operating characteristic curve [AUC-ROC] 0.803; **Fig. 5b**). Importantly, these models retained prognostic value when using PRO data from early monitoring periods. Using the first 30 days of PRO observations, the model achieved an AUC-ROC of 0.686, which improved with longer follow-up (60, 90, and 180 days: AUC-ROC 0.693, 0.726, and 0.78, respectively; **Fig. 5b**). These results indicate that PRO trajectories can provide clinically meaningful, early prediction of survival outcomes. By analyzing the feature importance in the machine learning predictive model, we identified that the PRO deterioration rates in pain/dyspnea and GI domains were the strongest predictors of survival. (**Fig. 5c**)

We also evaluated the prognostic value of PRO trajectory parameters derived from the total PRO score NLME model. Similar to symptom-level analyses, total PRO score trajectories demonstrated early predictive ability for OS, with AUCs of 0.71, 0.677, 0.716, and 0.745 using data from the first 30, 60, 90, and 180 days, respectively. Predictive performance was highest when using the full trajectory (AUC 0.79; **Fig. 5d**). Among all features, the rate of PRO deterioration in total scores was also the most important predictor of survival (**Fig. 5e**).

### PRO trajectories predict acute care utilization and treatment modifications

We examined whether PRO trajectory parameters could predict acute care utilization (hospitalization and ED visits) and treatment modifications (discontinuation and dose reduction). Using PRO data prior to event, the models achieved an AUC-ROC of 0.75 for hospitalization (**Fig. 6a**) and 0.65 for ED visits (**Fig. 6b**). PRO trajectories also predicted treatment discontinuation (AUC-ROC 0.707; **Fig. 6c**) and dose reduction (AUC-ROC 0.67; **Fig. 6d**).

**Figure 6.**
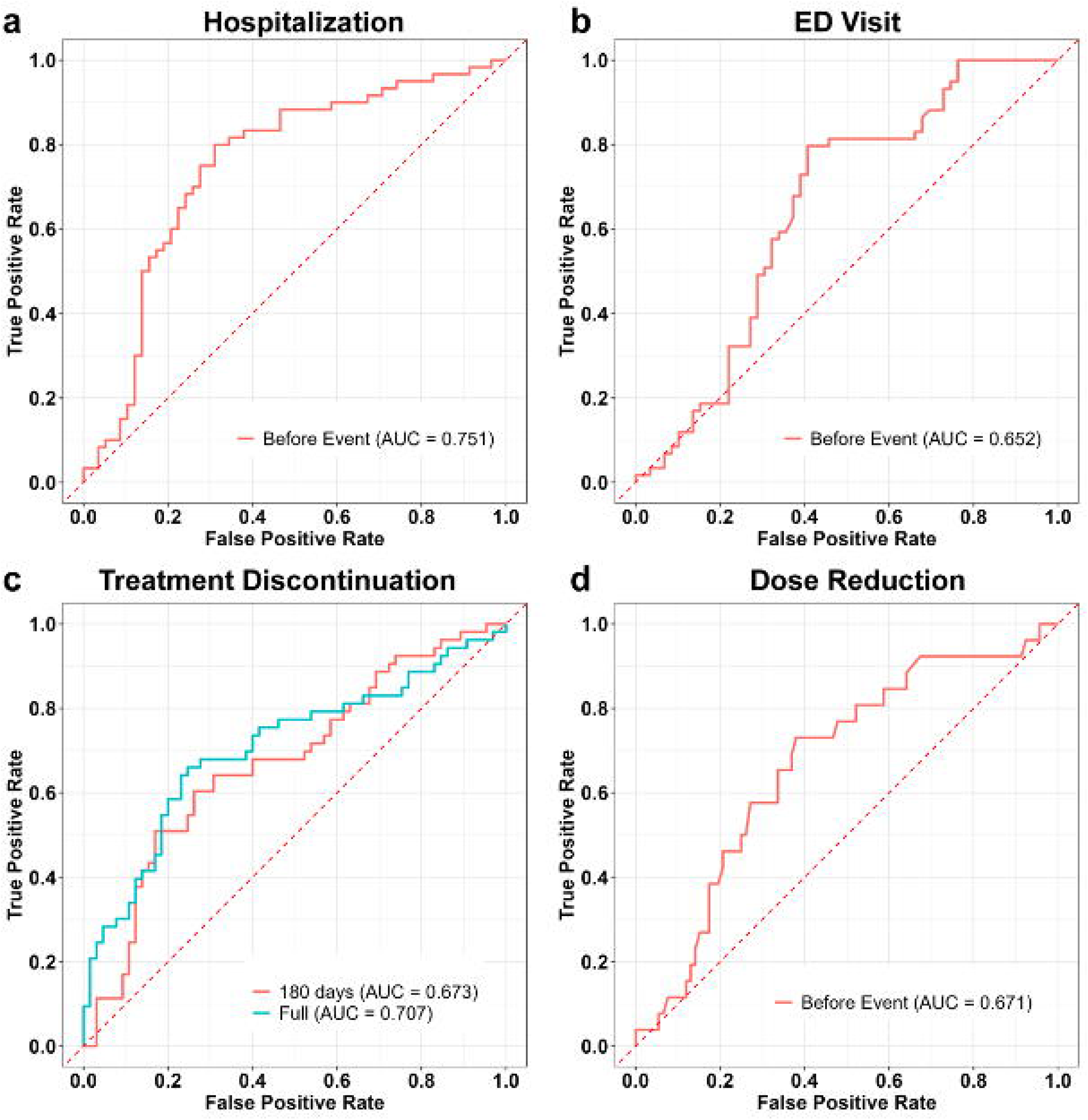
Patient-reported outcome (PRO) trajectories predict acute care utilization and treatment modification. (a–d) ROC curves for machine learning models predicting hospitalization (Random forest, a), ED visits (Gradient boosting, b), treatment discontinuation (Support vector machine, c), and dose reduction (Random forest, d). Model performance is summarized by the AUC. PRO, patient-reported outcome; ROC, Receiver operating characteristic; ED, emergency department; AUC, area under the curve.

Additional performance metrics, including accuracy, precision, recall, and F1-score, are provided in **Supplementary Table 6**. Together, these results further support the prognostic value of PRO trajectories for clinically relevant outcomes.

## Discussions

In this study, we developed a quantitative framework that integrates NLME modeling with IRT to characterize longitudinal PRO trajectories in patients with metastatic cancers. Using data from the PRO-TECT trial, we showed that both overall PRO scores and individual symptom trajectories can be reliably modeled over time. Symptom-level PROs provided greater clinical insight than total scores, revealing important differences across cancer types and treatments.

Importantly, PRO trajectory parameters were strongly associated with key clinical outcomes, including OS, acute care utilization, and treatment modifications. Models incorporating PRO trajectories were able to predict survival with good accuracy, even when using data from early observation periods. These findings support the use of PRO trajectories as clinically meaningful and actionable early indicators of patient outcomes.

Our modeling approach addresses a critical gap in oncology care: how to meaningfully integrate patients’ voices as clinically prognostic signals into decision-making. Although PRO data capture patients’ lived experiences during treatment, their use in routine practice remains limited, in part because clinicians lack clear frameworks to interpret and act on these data^18,19^. By transforming longitudinal PROs into quantitative trajectory parameters with demonstrated prognostic value, our approach provides a clinically interpretable link between patient-reported symptoms and key outcomes. This framework offers a practical pathway for incorporating PROs into risk stratification and care planning, which supports the adoption of more systematic and frequent PRO monitoring in routine oncology practice.

The IRT-NLME framework offers several unique advantages for analyzing longitudinal PRO data. First, it explicitly accounts for variability and noise, enabling separation of clinically meaningful symptom changes from random fluctuations. Second, it captures patient-specific PRO trajectories, allowing individualized risk prediction rather than relying on population averages. Third, by modeling symptom-level data, it preserves clinically relevant heterogeneity that is often obscured in composite PRO scores. Finally, despite 23.8% missingness, NLME model could leverage both individual-level information from prior to or after the missing value and population-level information to inform incomplete observations of some individuals.

Another key finding is that PRO data can support early prediction of adverse outcomes. Using only the first 3 months of PRO monitoring, the model predicted survival with an AUC-ROC of approximately 0.73, indicating good discrimination between patients who will survive and those at higher risk of death. More broadly, models based on PRO data collected within the first 30–180 days were able to provide informative predictions of future clinical events. These findings demonstrate that routinely collected PROs can serve as early warning signals, enabling clinicians to identify high-risk patients sooner and create opportunities for timely intervention, closer monitoring, and potential treatment modification.

Prior studies have applied several machine learning models to predict survival in cancer patients using PROs and have achieved relatively moderate performance (AUC-ROC between 0.66 to 0.76)^20–24^. However, most approaches focus on single-time-point PRO data, which provides only static snapshots of patient disease status, to predict survival. In contrast, our study leverages longitudinal PRO trajectories to characterize the dynamics of symptom progression.

Our model demonstrates more robust predictive performance for OS and preserves the ability to identify high-risk patients at earlier time points.

Several limitations should be considered. First, this study was based on a clinical trial, which may limit generalizability to real-world settings. External validation using real-world data is planned for future work. Second, the model did not consider the potential bidirectional relationship between PRO trajectories and clinical events. For example, treatment modifications or acute care utilization may in turn influence subsequent PROs and introduce potential confounding.

In summary, we present a quantitative framework that transforms longitudinal PRO data into clinically actionable predictors of patient outcomes. By integrating IRT and NLME modeling with machine learning, our approach converts patients’ symptom experiences into dynamic, patient-centered biomarkers with demonstrated prognostic value. Notably, these PRO-derived metrics enable prediction of survival even within the first months of monitoring. This work provides a practical pathway to incorporate patient-reported data into risk stratification and clinical decision-making. More broadly, it supports a shift toward patient-centered biomarkers that can complement traditional clinical measures and enhance precision oncology.

## Method

### Ethic Statement

The data used in this analysis are fully de-identified individual participant data for secondary analysis. The study was granted an Institutional Review Board (IRB) approval by the University of North Carolina at Chapel Hill.

### Trial Description

Data were collected from a multicenter cluster-randomized trial, PRO-TECT (ClinicalTrials.gov registration: NCT03249090). PRO-TECT trial enrolled patients with metastatic cancer who had previously received first-to fourth-line cancer therapy at the time of enrollment. The objective of this trial was to evaluate the impact of weekly electronic PRO symptom monitoring on clinical outcomes, compared with usual care group^4^. Our analysis is based on the PRO data from the PRO monitoring arm.

### Data Source

De-identified individual participant longitudinal PROs, baseline demographics and clinical characteristics, as well as clinical outcomes were included in the dataset.

PROs were assessed using the PRO-CTCAE, a National Cancer Institute (NCI) developed tool for patients to self-report the frequency, severity, and interference of symptomatic adverse events in cancer trials^3^. The full list of all PRO-CTCAE symptoms is shown in **Supplement Table S2**. Symptoms were grouped into three underlying diseases domains: gastrointestinal, psychological and pain and dyspnea. The PROs of each patient were collected weekly for one year or until voluntary withdrawal from the study or discontinuation of all cancer treatment. Considering the low frequency of level 4 responses in all symptoms, level 3 and 4 were grouped together in this analysis to ensure robust parameter estimation and sufficient sample size.

Baseline demographics and clinical characteristics include age, sex, race, ethnicity, cancer type, clinician-reported ECOG performance status, and prior intravenous and oral treatment of each patient. Clinical outcomes include overall survival (OS), emergency department (ED) visits, hospital admissions, dose reduction of the cancer drugs and treatment discontinuation.

### Model Development and Validation Workflow

The dataset was randomly split into a training cohort (80%) and a testing cohort (20%) for model development and validation. The overall analytical framework consisted of two sequential stages **(Fig 2a)**.

In **Stage 1**, longitudinal PRO data from the training cohort were used to derive PRO trajectory parameters for each individual patient. The performance of this step was evaluated by applying the trained model to PRO data from the testing cohort to ensure generalizability.

In **Stage 2**, the trajectory parameters generated from Stage 1 were used as inputs to train machine learning models for predicting clinical outcomes, including OS, hospitalization, ED visits, treatment discontinuation, and dose reduction. Model performance was then evaluated in the testing cohort using trajectory parameters derived from the testing data.

To further assess early prediction capability, we conducted additional analyses using truncated PRO data from the testing cohort. Specifically, trajectory parameters were estimated using PRO data collected within the first 30, 60, 90, and 180 days. These parameters were then input into the trained machine learning models to evaluate predictive performance based on early observation windows.

### Nonlinear Mixed Effects (NLME) Modeling Approach

To characterize longitudinal PRO trajectories over time, we applied NLME modeling approach^7,8^. The model structure was a semi-mechanistic model (**Fig. 2b**), which describes symptom improvement using an asymptotic exponential function and symptom worsening using a linear deterioration component. The NLME approach allows us to estimate PRO trajectory parameters at both the population and individual levels. Specifically, the model separates the underlying PRO trajectory from variability and noise by quantifying (1) the average trajectory across all patients, (2) how individual patients deviate from this average pattern, and (3) residual variability due to unexplained random fluctuations (measurement noise) (**Fig. 2c**). Model parameters were estimated using all available longitudinal PRO data through a likelihood-based framework implemented with the Stochastic Approximation Expectation-Maximization (SAEM) algorithm^25^. This method enables robust estimation of both population-level parameters and patient-specific parameters, even in the presence of irregular and sparse observations. In this analysis, we built two NLME models: (1) a beta-regression NLME model based on total PRO scores, and (2) an item response theory (IRT) embedded NLME model using individual symptom scores.

Additional details on the model structure, assumptions, and estimation procedures are provided in **Supplementary Materials**.

### Machine Learning Models Predicting Clinical Outcomes

To evaluate the predictive value of PROs, machine learning algorithms were used to predict clinical outcomes using NLME-derived PRO trajectory parameters as input features. Several algorithms were tested, including logistic regression, random forest, gradient boosting, support vector machine and XGboost. Model performance was primarily assessed using the area under the receiver operating characteristic curve (AUC-ROC), which quantifies the model’s ability to distinguish between patients with or without specific clinical outcomes^26^. Other metrics include accuracy, precision, sensitivity, F1-score were also examined. To ensure fair comparisons across models, we (1) used identical training and test cohort (80% for training data cohort and 20% for testing data cohort), (2) applied consistent preprocessing pipelines (including scaling, missing value imputation, and feature encoding), and (3) optimized hyperparameters using the same search strategy with cross-validation performed in the training cohort.

### Missing Data Handling

For the beta-regression NLME model, missing PRO scores were imputed using the last observation carried forward (LOCF) approach. For the IRT-NLME model, missing scores were excluded from the analysis and no further imputation was performed.

The missingness for all covariates was low (<1%). Therefore, simple imputation methods were applied, with continuous variables imputed using the median and categorical variables imputed using the mode.

## Statistical Analysis

Multivariable logistic regression was used to evaluate associations between baseline covariates and clinical outcomes (dose reduction, hospitalization, treatment discontinuation and ED visits). Odds ratios (ORs), 95% confidence intervals (CIs), and p-values were reported. Time-to-event data (Survival) was analyzed using multivariable Cox proportional hazards models. Results were presented as hazard ratios (HRs) with 95% CIs and visualized by forest plots. Statistical significances were defined as p < 0.05. In post hoc analyses, univariate logistic regression models were constructed to evaluate the associations between IRT derived individual parameters and clinical outcomes, and the results were visualized using a heatmap. Parameter distributions comparison across cancer types and treatment groups were evaluated using the Kruskal–Wallis’s test, followed by Dunn’s test for pairwise comparisons. P-values were adjusted using the Holm method, with adjusted p < 0.05 considered statistically significant.

## Data Availability

Raw datasets are available upon reasonable request from corresponding author for secondary data analyses. Two separate synthetic datasets are provided to allow readers to test and run the model codes for beta-regression NLME model and the IRT-based NLME model, respectively. These datasets include the key model inputs of the original data and do not contain any identifiable patient information.

## Analysis Codes Availability

Data cleaning, statistical analyses and data visualization were performed using R 4.4.1 and RStudio Version 2025.05.13+513. NLME models were developed using Monolix 2024R1 and model simulations were performed using Simulx 2024R1. Both Monolix and Simulx could be downloaded at https://lixoft.com/products/. The machine learning analyses were conducted in Python 3.12 using scikit-learn, XGBoost, pandas, and NumPy packages. The figures were compiled in Adobe Illustrator 2025. The source codes were provided in GitHub (https://github.com/zhangcy000/PRO-TECH-Trial).

## Data Availability

All data generated in this study are available from the authors upon reasonable request. To support reproducibility of the methods, synthetic datasets are provided for readers to use in testing the models.

https://github.com/zhangcy000/PRO-TECH-Trial

## Acknowledgements

The authors thank the patients and their families and caregivers and investigators for participating in the studies used in this analysis. We acknowledged the use of AI tools (OpenAI, ChatGPT) to improve the language in the manuscript writing.

