## Supplementary Materials for "Transforming Patient Voices into Early Predictors of Survival Using Nonlinear Mixed-Effect Models and AI/ML for Patient-Centered Decision-Making"

#### Supplementary Methods

#### 1.1 Bounded data beta-transformation model for PRO total score

Longitudinal trajectories of total PRO scores, aggregated across all 14 symptoms, were characterized by a semi-mechanistic model that captured both treatment-related improvement and toxicity-related deterioration **(Fig 2b)**. The improvement was modeled using an asymptotic exponent function, characterized by Emax (maximum efficacy) and $K_d$ (improvement rate). The deterioration was described as a linear function with a rate constant (*SLP*). The PRO score trajectory model is shown below:

$$PRO_i(t) = PRO_{i,base} - E_{max} \times (1 - e^{-K_d t}) + SLP \times 0.001 \times t \quad \text{(Equation 1)}$$

where $PRO_{i,j}(t)$ is PRO score at time *t* in days for subject *i* and $PRO_{i,base}$ is the PRO score at baseline for subject *i*.

Because many PRO responses cluster at the scale boundaries (0 or 4), the total PRO scores also exhibit a high frequency of boundary values (0 and 56). To account for these boundary values, zero-/one-inflated beta regression equations were applied to characterize the data[1]. The PRO scores were treated as discrete integer variables bounded between 0 to 56 in this analysis.

$$p(y_{ij}) = \begin{cases} p0 & if\ y_{ij} = 0 \\ p1 & if\ y_{ij} = 1 \\ (1 - p0 - p1)\frac{\Gamma(\tau)}{\Gamma(\mu_{ij}\tau)\Gamma((1-\mu_{ij})\tau)} {y_{ij}}^{(\mu_{ij}\tau-1)}(1 - y_{ij})^{(1-\mu_{ij})\tau-1} & otherwise \end{cases}$$

(Equation 2)

Where $y_{ij}$ is the PRO score reported by subject $i$ at time $j$, scaled to a value between 0 and 1. The beta distribution component is defined using the gamma function ($\Gamma$). The parameter $\mu_{ij}$ represents the expected value of $y_{ij}$, while $\tau$ is the precision parameter that determines how tightly the values cluster around the mean in the gamma function.

To model the probabilities of boundary values ($p0$ and $p1$), we used logistic functions based on the logit transformation of the central value $\mu_{ij}$:

$$\mu_{ij} = \frac{exp(PRO_i(t))}{1+exp\ (PRO_i(t))} \quad \text{(Equation 3)}$$

$$logit(p0) = \ -\gamma 0 - \gamma 1 \times \log\left(\frac{\mu_{ij}}{1-\ \mu_{ij}}\right) \quad \text{(Equation 4)}$$

$$logit(p1) = \ -\gamma 0 + \gamma 1 \times \log\left(\frac{\mu_{ij}}{1-\ \mu_{ij}}\right) \quad \text{(Equation 5)}$$

Where $\gamma 0$ and $\gamma 1$ are the intercept and slope parameters in the beta-transformation that control how the probabilities of observing PRO scores at the boundaries vary with the expected value $\mu_{ij}$.

#### 1.2 Item-response theory (IRT) model for individual symptoms

The IRT model was developed in two steps: (1) a base IRT model was constructed to link the probability of each symptom response to underlying disease variable; (2) a longitudinal structure model was built to describe the trajectory of this underlying disease variable over time. **(Fig 3b)**

*Base IRT Model*

The IRT describes the probability of each response level for a given symptom as a function of patients' underlying disease variable ($L$), representing patient status[2,3]. Our IRT model included three of these variables to account for the three symptoms domains (**Fig 3a**). The graded response model was used to characterize each symptom[3,4], where probability for subject $i$ to have at least a response of $s$ (s = 0 to 3) for symptom $j$ was described in the equation 6 and the probability of having the score s is calculated based on equation 7.

$$P(y_{ij} \geq s) = \frac{1}{1+e^{-a_j(L_{i,j}-b_{j,s})}} \quad \text{(Equation 6)}$$

$$P(y_{ij} = s) = P(y_{ij} \geq s) - P(y_{ij} \geq s+1) \quad \text{(Equation 7)}$$

where $a_j$ is the discrimination parameter of the symptom and $b_{j,s}$ is the difficulty parameter for the response of level s of the symptom $j$. $b_{j,s}$ are non-decreasing for the same symptoms ($b_{j,s+1} > b_{j,s}$). $L_{i,j}$ represents the variable for symptom domain $j$ of patient $i$. Higher $L_{i,j}$ value denotes less symptom burden (or patient feel better) for symptom domain $j$. The PRO score was reversed coded as 3, 2, 1 and 0 (e.g., the higher the score the lower the symptom burden). For example, $P(y_{ij} = 3)$ means the reported PRO is 0. $L_{i,j}$ was assumed to be a normal distribution with mean $L_j$ and variance 1 at baseline and estimated variance in the later longitudinal model. Item characteristic curves (ICC) of each symptom were generated to show the probability of each response level for a given level of patient underlying disease variable ($L$). The shape of the ICCs is determined by $a_j$ and $b_{j,s}$.

*Longitudinal IRT Model*

A similar equation as the previous beta-regression model was used to characterize the longitudinal changes of the underlying disease status for each symptom domain of each subject:

$$L_{i,j}(t) = L_{i,j,BASE} + E_{\max_{i,j}} \times \left(1 - e^{-K_{d_{i,j}}t}\right) - SLP_{i,j} \times 0.001 \times t \quad \text{(Equation 8)}$$

where $L_{i,j}(t)$ is underlying disease status for symptom domain $j$ at time $t$ in days for subject $i$ and $L_{i,j,BASE}$ is the underlying disease status at baseline for symptom domain $j$ for subject $i$. $E_{\max_{i,j}}$ is the maximum symptom improvement for symptom domain $j$ for subject $i$. $K_{d_{i,j}}$ and $SLP_{i,j}$ are the symptom improvement and deterioration rates for symptom domain $j$ for subject $i$.

#### 1.3 NLME model development

To explain the heterogeneity across subjects, inter-individual variability (IIV) was incorporated into the NLME model parameters. The IIV was described using a normal or log-normal distribution, as defined in equations (9) and (10) respectively

$P_i = \theta_P + \eta_i$ (Equation 9)

$P_i = \theta_P \cdot e^{\eta_i}$ (Equation 10)

Where $P_i$ represents the individual parameter value for subject $i$, $\theta_P$ is the typical population value of parameter $P$, and $\eta_i$ is a random effect assumed to follow a normal distribution with mean 0 and variance $\omega^2$. For the beta-regression NLME model, IIV was assumed to be a normal distribution for parameter $E_{max}$, $PRO_{base}$ and $SLP$ and a log-normal distribution for parameter $K_d$ . For IRT NLME model, IIV was assumed to be a normal distribution for parameter $L_{BASE}$, and a log-normal distribution for parameter $E_{max}, K_d$ and $SLP$.

#### 1.4 Covariate model development

A covariate model was developed to evaluate the effect of the potential baseline covariates on relevant model parameters based on exploratory analysis and clinical relevance. Investigated baseline covariates included patient demographics (sex, age, race and ethnicity), and Eastern Cooperative Oncology Group (ECOG) performance status. Cancer types and treatments were not included in the covariate model but were evaluated in post hoc analyses. Covariate analysis was performed using the stepwise covariate model building procedure (SCM), with a significance level of $p < 0.01$ in the forward selection and $p < 0.001$ in the backward elimination.

The effect of a categorical covariate on a parameter was described as a linear relationship as shown in equation (5) and (6). Centered linear relationships were used to describe the effect of a continuous covariate on a parameter as shown in equation (7).

$$P_i = \theta_P + \theta_{COV} \quad \text{(Equation 11)}$$

$$\theta_{COV} = \begin{cases} 0 \quad if\ reference\ category \\ \theta_{COV,1} \qquad if\ category\ 1 \\ \theta_{COV,2} \qquad if\ category\ 2 \\ \qquad\qquad \ldots \end{cases} \quad \text{(Equation 12)}$$

$$P_i = \theta_P + \theta_{COV}(COV_i - COV_{median}) \quad \text{(Equation 13)}$$

where $P_i$ is the individual parameter value, $\theta_P$ is the typical value of the parameter $P$, $\theta_{COV}$ is the coefficient for the effect of the covariate on parameter $P$. $COV_i$ is the continuous covariate value for subject $i$ and $COV_{median}$ is the median value of the covariate in the population. Missing values for covariates were imputed with the population median (for continuous covariates) or mode (for categorical covariates) during the covariate analysis. The impact of baseline covariates on latent PRO score from beta-regression model was evaluated using model-based simulation. A total of 1000 simulations were conducted to display the effect of covariate on parameter distributions.

**1.5 Model selection**

Model selection was based on the minimum objective function value (OFV), diagnostic plots and the precisions of parameter estimations. For nested models, a decrease in OFV of ≥ 6.63 ($p < 0.01$) was considered statistically significant improvement. The predictive performance of the models was assessed through visual predictive checks (VPCs). VPC is a tool in NLME models to compare observed data to model-simulated prediction intervals to assess the model's ability to capture the central tendency and variability of the data[5]. A total of 200 Monte Carlo simulations of the dataset were performed to generate the VPCs. Due to the substantial drop-out in the trial and may cause bias in the VPC, we corrected the VPC for dropout by incorporating dropout predictions into the model and excluding individual predictions occurring after the corresponding predicted dropout time[6]. Parameter estimations were assessed by two metrics: (1) Relative standard error (RSE) reflects the precision of parameter estimates. Generally, RSE values below 30% indicate stable and reliable parameter estimation. (2) Shrinkage is the parameter indicating how much individual parameter estimates were pulled towards population typical parameter values. Shrinkage < 40% suggests the model captures sufficient between-subject variability. A high shrinkage (> 50%) means the individual predictions may not be reliable—even when model diagnostics look acceptable overall.

### Supplementary Figures

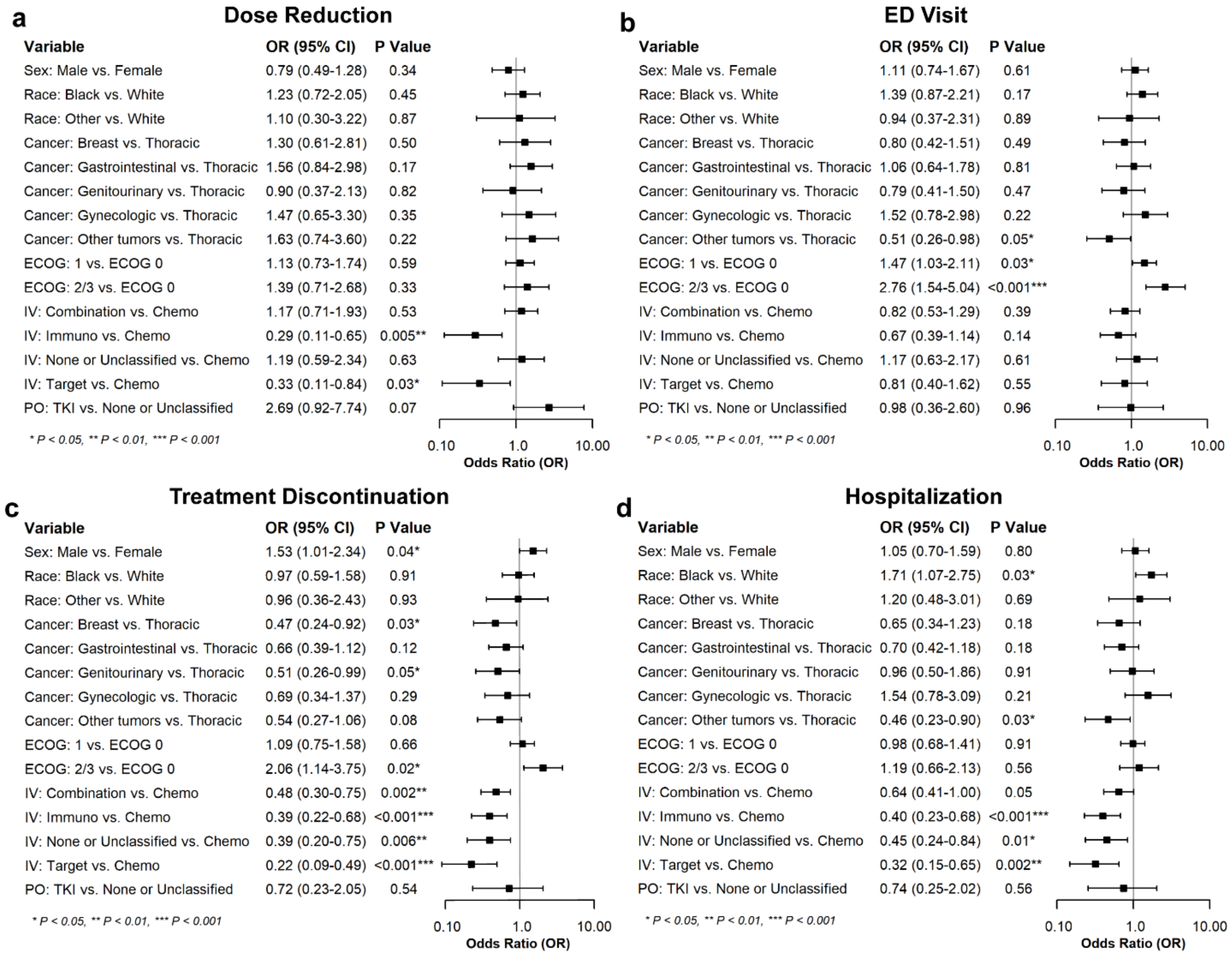

**Supplement Figure 1: Multivariable logistic regression of baseline characteristics with clinical outcomes.**

(a-d) Forest Plot for baseline demographics with clinical outcomes. (a) Dose Reduction; (b) ED visit; (c) Treatment discontinuation; (d) Hospitalization event. ED, emergency department; IV, intravenous treatment; PO, oral treatment; TKI, tyrosine kinase inhibitors.

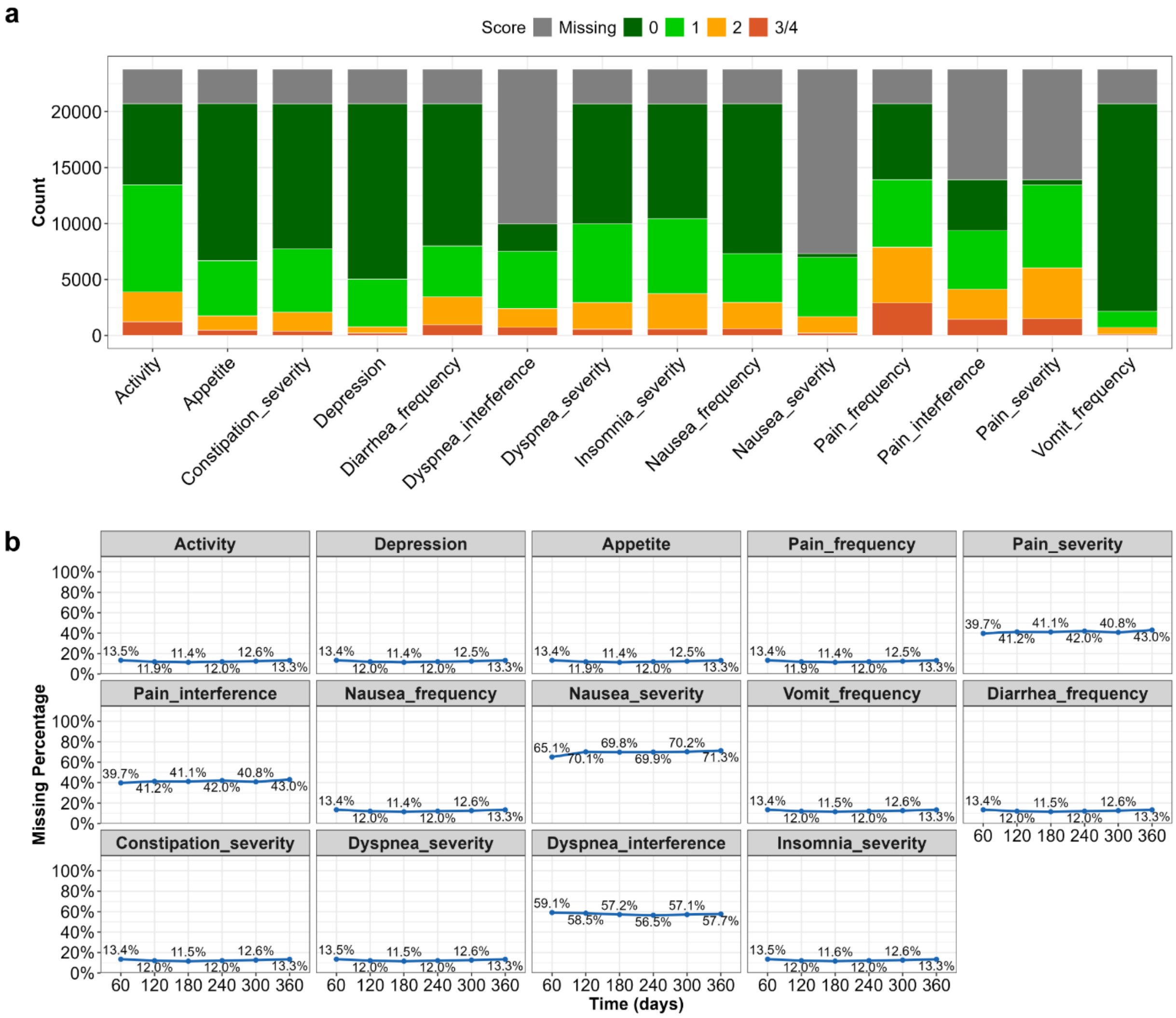

**Supplement Figure 2: Distribution and missingness of the PRO data.**

(a) Distribution of observed PRO- levels across all symptoms. (b) Missing percentage of each PRO symptom. Missingness percentages were summarized within each 60-day time window. PRO, Patient-Reported Outcomes.

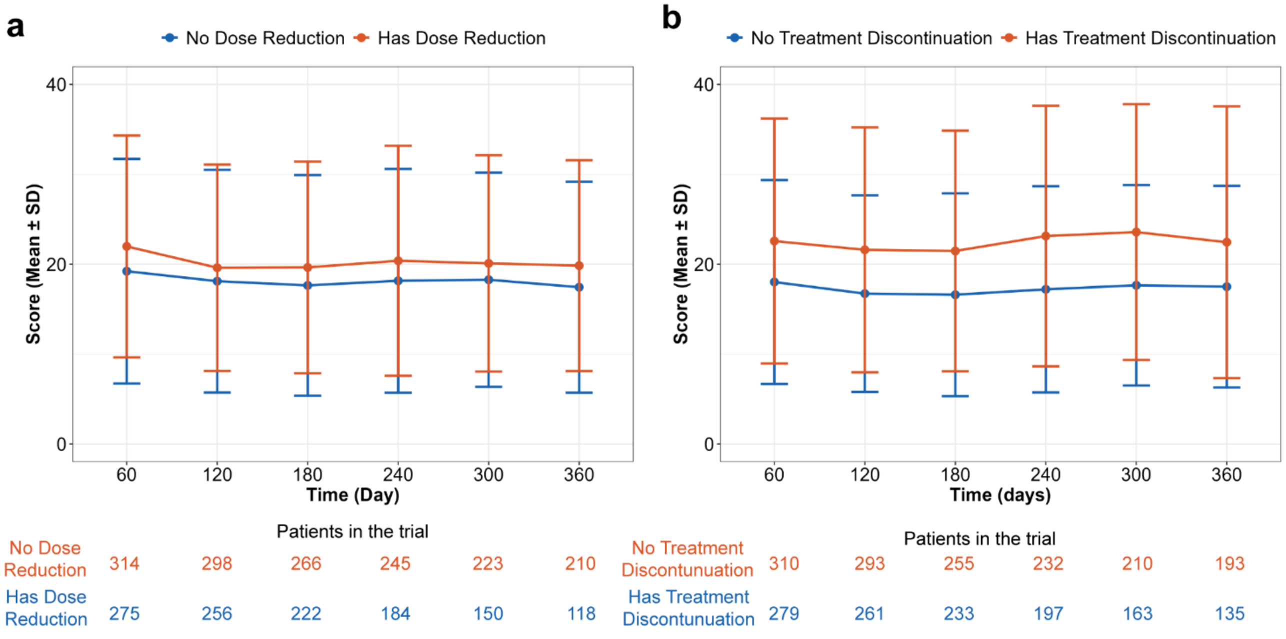

**Supplement Figure S3: Longitudinal patient-reported outcome (PRO) scores in the PRO-TECT trial.**

(a) Longitudinal PRO total scores (mean ± s.d.) in patients with and without dose reduction. (b) Longitudinal PRO total scores (mean ± s.d.) in patients with and without treatment discontinuation. s.d., standard deviation.

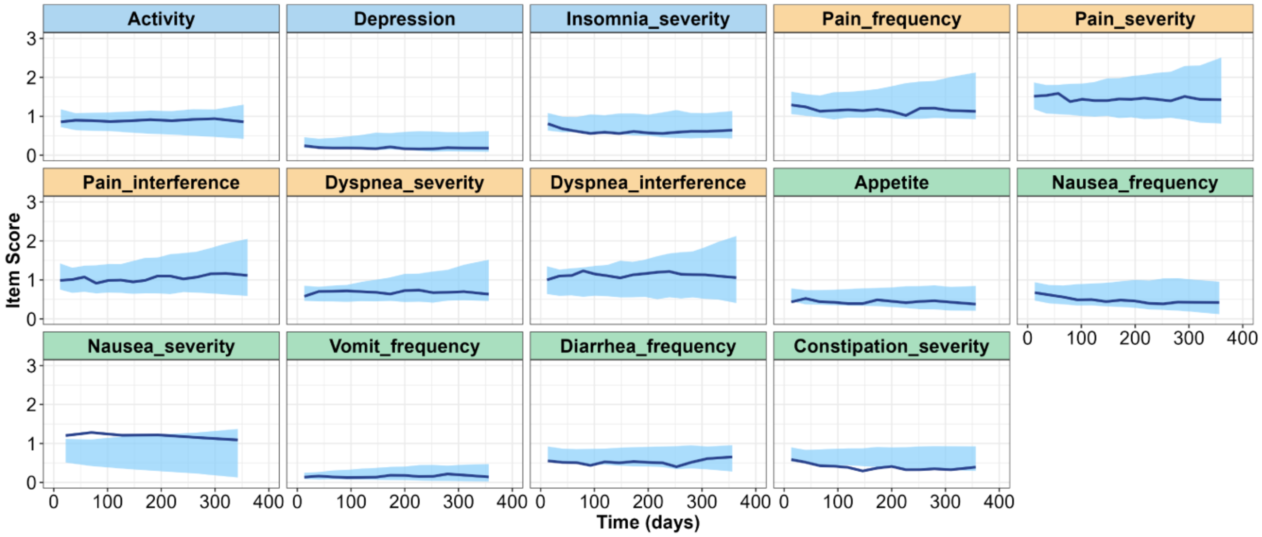

**Supplement Figure S4: Visual predictive check (VPC) of patient-reported outcome (PRO) trajectories using full testing cohort.**

VPC evaluates the performance of IRT-NLME model with the full longitudinal trajectories of the testing cohort. Solid lines indicate the median observed PRO total scores over time, and shaded areas represent the 95% prediction interval from the model. Good agreement between the shaded regions and the observed data indicates that the model adequately captures the underlying data patterns. NLME, nonlinear mixed-effects; IRT, item-response theory. Panels' color corresponds to the symptom domain: blue for psychological, orange for pain/dyspnea, and green for gastrointestinal.

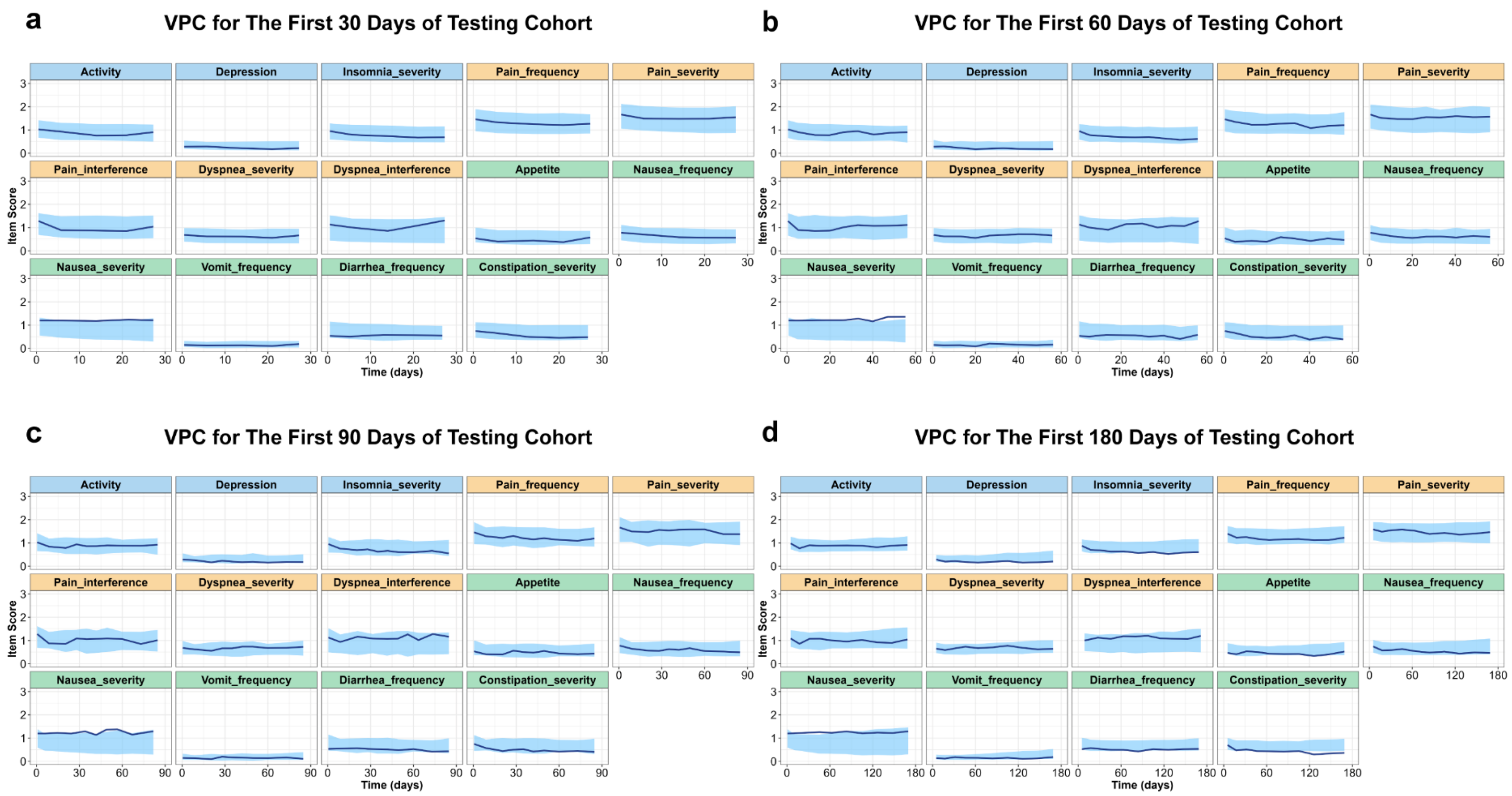

**Supplement Figure S5: Visual predictive check (VPC) of patient-reported outcome (PRO) trajectories using early observations of the testing cohort.**

(a-d) VPC evaluates the performance NLME model with the IRT extension in testing cohort. (a) First 30 days; (b) First 60 days; (c) First 90 days; (d) First 180 days. Solid lines indicate the median observed PRO total scores over time, and shaded areas represent the 95% prediction interval from the model. Good agreement between the shaded regions and the observed data indicates that the model adequately captures the underlying data patterns. NLME, nonlinear mixed-effects; IRT, item-response theory. Panels' color corresponds to the symptom domain: blue for psychological, orange for pain/dyspnea, and green for gastrointestinal.

### Supplementary Tables

#### Supplement Table S1: Clinical Endpoint Time

| Event Type | Time to First Event, Median (Range), days | Training Dataset (N = 471) | | Testing Dataset (N = 118) | | Whole Dataset (N = 589) | |
|---|---|---|---|---|---|---|---|
| | | No | Yes | No | Yes | No | Yes |
| Dose Reduction | 77 (0 – 338) | 360 (76.4%) | 111 (23.6%) | 92 (78%) | 26 (22%) | 452 (76.7%) | 137 (23.3%) |
| Treatment Discontinuation | NA | 289 (61.4%) | 182 (38.6%) | 65 (55%) | 53 (45%) | 354 (60.1%) | 235 (39.9%) |
| ED Visit | 100.5 (2 – 434) | 251 (53.3%) | 220 (46.7%) | 59 (50%) | 59 (50%) | 310 (52.6%) | 279 (47.4%) |
| Hospitalization | 108 (3 – 400) | 256 (54.4%) | 215 (45.6%) | 58 (49.2%) | 60 (50.8%) | 314 (53.3%) | 275 (46.7%) |
| Survival Time | 303.5 (32 – 739) | 204 (43.3%) | 267 (56.7%) | 47 (39.8%) | 71 (60.2%) | 251 (42.6%) | 338 (57.4%) |

Note: Patients without available time were excluded from the median calculation.

**Supplement Table S2: PRO Items and Response Category Definitions**

| Symptoms Domain | Variable Name | Question Label | Category Level | | | | |
|---|---|---|---|---|---|---|---|
| | | | 0 | 1 | 2 | 3 | 4 |
| Gastrointestinal | Appetite_overall | In the last 7 days, has your EATING or DRINKING DECREASED? | “Not at all” | “A little bit” | “Somewhat” | “Quite a bit” | “Very much” |
| | PROCTCAE_Nausea_f | In the last 7 days, how OFTEN did you have NAUSEA? | “Never” | “Rarely” | “Occasionally” | “Frequently” | “Almost constantly” |
| | PROCTCAE_Nausea_s | In the last 7 days, what was the SEVERITY of your NAUSEA at its WORST? | “None” | “Mild” | “Moderate” | “Severe” | “Very Severe” |
| | PROCTCAE_Vomit_f | In the last 7 days, how OFTEN did you have VOMITING? | “Never” | “Rarely” | “Occasionally” | “Frequently” | “Almost constantly” |
| | PROCTCAE_Diarr_f | In the last 7 days, how OFTEN did you have LOOSE OR WATERY STOOLS (DIARRHEA)? | “Never” | “Rarely” | “Occasionally” | “Frequently” | “Almost constantly” |
| | PROCTCAE_Constip_s | In the last 7 days, what was the SEVERITY of your CONSTIPATION at its WORST? | “None” | “Mild” | “Moderate” | “Severe” | “Very Severe” |
| Psychological | Activity | In the last 7 days, how would you generally rate your ACTIVITY: | "Normal with no limitations” | "Not your normal self, but able to be up and about with fairly normal activities" | "Not feeling up to most things, but in bed or chair less than half the day" | "Able to do little activity & spend most of the day in bed or chair" | “Pretty much bedridden, rarely out of bed" |
| | Depression | In the last 7 days, how often have you been bothered by FEELING DOWN, DEPRESSED, OR HOPELESS? | "Not at all" | "Several days" | "More than half the days" | "Nearly every day" | NA |

| | | | | | | | |
|---|---|---|---|---|---|---|---|
| | PROCTCAE_Insomnia_s | In the last 7 days, what was the SEVERITY of your INSOMNIA (including difficulty falling asleep, staying asleep, or waking up too early) at its WORST? | "None" | "Mild" | "Moderate" | "Severe" | "Very Severe" |
| Pain and Dyspnea | PROCTCAE_Pain_f | In the last 7 days, how OFTEN did you have PAIN? | "Never" | "Rarely" | "Occasionally" | "Frequently" | "Almost constantly" |
| | PROCTCAE_Pain_s | In the last 7 days, what was the SEVERITY of your PAIN at its WORST? | "None" | "Mild" | "Moderate" | "Severe" | "Very Severe" |
| | PROCTCAE_Pain_i | In the last 7 days, what was the INTERFERENCE of your PAIN at its WORST? | "Not at all" | "A little bit" | "Somewhat" | "Quite a bit" | "Very much" |
| | PROCTCAE_Dyspnea_s | In the last 7 days, what was the SEVERITY of your SHORTNESS OF BREATH at its WORST? | "None" | "Mild" | "Moderate" | "Severe" | "Very Severe" |
| | PROCTCAE_Dyspnea_i | In the last 7 days, what was the INTERFERENCE of your SHORTNESS OF BREATH at its WORST? | Not at all | A little bit | Somewhat | Quite a bit | Very much |

**Supplement Table S3: Final PRO Score Beta-regression model parameter estimates.**

| Parameter | Value | RSE (%) | SHR (%) |
|---|---|---|---|
| **Population Parameters** | | | |
| Baseline Patient-reported Outcomes Score ($BASEP$)* | -1.8 | 4.1 | 3.65 |
| Maximal Symptom improvements ($EMAX$)* | 0.53 | 11.1 | 13.6 |
| Symptom improvement rate ($KD$, 1/day)* | 0.013 | 6.75 | 45.0 |
| Symptom deterioration rate ($SLP$, 1/day)* | 2.15 | 10.5 | 15.6 |
| Beta-transformation precision (τ) | 54.57 | 1.08 | / |
| Beta-transformation slope (γ0) | 23.53 | 6.98 | / |
| Beta-transformation intercept (γ1) | 6.32 | 7.71 | / |
| **Inter-individual variability (standard deviation)** | | | |
| Standard deviation of $BASEP$ ($\Omega_{BSV\_BASE}$) | 0.75 | 3.45 | |
| Standard deviation of $EMAX$ ($\Omega_{BSV\_EMAX}$) | 1.14 | 3.98 | |
| Standard deviation of $KD$ ($\Omega_{BSV\_KD}$) | 0.93 | 6.02 | |
| Standard deviation of $SLP$ ($\Omega_{BSV\_SLP}$) | 4.35 | 4.38 | |
| **Covariates** | | | |
| ECOG I on BASEP* | 0.27 | 28.6 | |
| ECOG II/III on BASEP* | 0.56 | 21.8 | |
| Female on BASEP* | 0.3 | 24.9 | |

*Parameter were beta-transformed.

**Supplement Table S4: Item-specific parameter estimates from the final longitudinal item-response theory model.**

| Item | a | $b_1$ | $\delta_2$ | $\delta_3$ |
|---|---|---|---|---|
| Activity | 1.56 | 0.96 | 0.99 | 1.71 |
| Depression | 2.12 | 0 | 0.9 | 1.32 |
| Appetite | 1.14 | 1.51 | 1.05 | 1.27 |
| Frequency of Pain | 1.0 | 1.12 | 1.5 | 1.14 |
| Severity of Pain | 1.0 | 0.41 | 2.18 | 3.13 |
| Interference of Pain | 1.0 | 0.49 | 1.23 | 1.51 |
| Frequency of Nausea | 4.17 | 2.68 | 0.85 | 0.55 |
| Severity of Nausea | 4.97 | 2.25 | 0.99 | 1.44 |
| Frequency of Vomit | 1.75 | 0 | 1.98 | 0.69 |
| Frequency of Diarrhea | 0.6 | 0 | 2.1 | 1.69 |
| Severity of Constipation | 0.65 | 0 | 1.94 | 2.11 |
| Severity of Dyspnea | 0.9 | -1.46 | 1.95 | 1.95 |
| Interference of Dyspnea | 1.35 | 0 | 1.32 | 1.87 |
| Severity of Insomnia | 1.02 | 0 | 1.88 | 1.4 |

$b_2 = b_1 + \delta_2$; $b_3 = b_2 + \delta_3$

**Supplement Table S5: Final Item - Response Theory model parameter estimates.**

| Parameter | Value | RSE (%) | SHR (%) |
|---|---|---|---|
| **Population Parameters** | | | |
| Baseline Gastrointestinal Symptoms (*BASEGI*) | 4.18 | 1.5 | 10.5 |
| Baseline Psychological Symptom (*BASEPS*) | 3.33 | 2.9 | 10.9 |
| Baseline Pain and Dyspnea Symptom (*BASEPD*) | 2.93 | 3.7 | 8.4 |
| Maximal Gastrointestinal Symptom improvements (*EMAXGI*) | 1.61 | 5.7 | 36.9 |
| Maximal Psychological Symptom improvements (*EMAXPS*) | 1.18 | 6.8 | 43.5 |
| Maximal Pain and Dyspnea Symptom improvements (*EMAXPD*) | 1.58 | 7.8 | 44.3 |
| Gastrointestinal Symptom improvement rate (*KDGI*, 1/day) | 0.0051 | 12.2 | 34.4 |
| Psychological Symptom improvement rate (*KDPS*, 1/day) | 0.004 | 14.6 | 41.7 |
| Pain and Dyspnea Symptom improvement rate (*KDPD*, 1/day) | 0.0041 | 13.2 | 40.5 |
| Gastrointestinal Symptom deterioration rate (*SLPGI*, 1/day) | 1.4 | 10.2 | 30.7 |
| Psychological Symptom deterioration rate (*SLPPS*, 1/day) | 1.49 | 9.4 | 33.3 |
| Pain and Dyspnea Symptom deterioration rate (*SLPPD*, 1/day) | 2.48 | 8.8 | 26.9 |
| **Inter-individual variability (standard deviation)** | | | |
| Standard deviation of *BASEGI* ($\Omega_{BSV\_BASEGI}$) | 0.79 | 3.76 | |
| Standard deviation of *BASEPS* ($\Omega_{BSV\_BASEPS}$) | 0.96 | 4.66 | |
| Standard deviation of *BASEPD* ($\Omega_{BSV\_BASEPD}$) | 1.49 | 3.84 | |
| Standard deviation of *EMAXGI* ($\Omega_{BSV\_EMAXGI}$) | 0.99 | 4.66 | |
| Standard deviation of *EMAXPS* ($\Omega_{BSV\_EMAXPS}$) | 0.99 | 5.38 | |
| Standard deviation of *EMAXPD* ($\Omega_{BSV\_EMAXPD}$) | 1.16 | 5.61 | |
| Standard deviation of *KDGI* ($\Omega_{BSV\_KDGI}$) | 2.21 | 4.6 | |
| Standard deviation of *KDPS* ($\Omega_{BSV\_KDPS}$) | 2.22 | 6.84 | |
| Standard deviation of *KDPD* ($\Omega_{BSV\_KDPD}$) | 1.94 | 6.07 | |
| Standard deviation of *SLPGI* ($\Omega_{BSV\_SLPGI}$) | 1.6 | 4.96 | |
| Standard deviation of *SLPPS* ($\Omega_{BSV\_SLPPS}$) | 1.38 | 5.16 | |

| | | |
|---|---|---|
| Standard deviation of *SLPPD* ($\Omega_{BSV\_SLPPD}$) | 1.37 | 5.34 |
| **Covariates** | | |
| ECOG I on BASEPS | -0.47 | 21.1 |
| ECOG II/III on BASEPS | -0.99 | 16.0 |
| ECOG I on BASEPD | -0.51 | 29.7 |
| ECOG II/III on BASEPD | -1.18 | 20.4 |
| Female on BASEGI | -0.33 | 24.1 |
| Female on BASEPS | -0.37 | 26.1 |

RSE, relative standard error; SHR, shrinkage.

**Supplement Table S6: Machine learning model performance for different clinical outcomes.**

| Clinical Endpoint | Machine Learning Model | Testing Data Range | Accuracy | Macro Average | | |
|---|---|---|---|---|---|---|
| | | | | Precision | Recall | F1-Score |
| Treatment Discontinuation – IRT model | Support vector machine | 30 days | 0.58 | 0.55 | 0.30 | 0.39 |
| | | 60 days | 0.50 | 0.47 | 0.96 | 0.63 |
| | | 90 days | 0.58 | 0.53 | 0.60 | 0.57 |
| | | 180 days | 0.68 | 0.65 | 0.60 | 0.63 |
| | | **Full** | **0.71** | **0.69** | **0.66** | **0.67** |
| Dose Reduction – IRT model | Random Forest | 30 days | 0.65 | 0.33 | 0.58 | 0.42 |
| | | 60 days | 0.77 | 0.48 | 0.54 | 0.51 |
| | | 90 days | 0.50 | 0.28 | 0.81 | 0.42 |
| | | **Before Event** | **0.64** | **0.35** | **0.73** | **0.48** |
| Hospitalization – IRT model | Random Forest | 30 days | 0.55 | 0.54 | 0.83 | 0.65 |
| | | 60 days | 0.59 | 0.73 | 0.32 | 0.44 |
| | | 90 days | 0.60 | 0.70 | 0.38 | 0.49 |
| | | **Before Event** | **0.75** | **0.73** | **0.80** | **0.76** |
| ED Visit – IRT model | Gradient boosting | 30 days | 0.57 | 0.55 | 0.81 | 0.65 |
| | | 60 days | 0.59 | 0.61 | 0.53 | 0.56 |
| | | 90 days | 0.60 | 0.60 | 0.63 | 0.61 |
| | | **Before Event** | **0.69** | **0.66** | **0.80** | **0.72** |
| Overall Survival – IRT model | Logistic Regression | 30 days | 0.71 | 0.71 | 0.87 | 0.78 |
| | | 60 days | 0.69 | 0.76 | 0.72 | 0.74 |
| | | 90 days | 0.75 | 0.74 | 0.89 | 0.81 |
| | | 180 days | 0.71 | 0.82 | 0.66 | 0.73 |
| | | **Full** | **0.78** | **0.76** | **0.92** | **0.83** |
| Overall Survival – PRO total model | Logistic Regression | 30 days | 0.71 | 0.74 | 0.80 | 0.77 |
| | | 60 days | 0.68 | 0.77 | 0.66 | 0.71 |
| | | 90 days | 0.69 | 0.77 | 0.70 | 0.74 |
| | | 180 days | 0.68 | 0.87 | 0.55 | 0.67 |
| | | **Full** | **0.75** | **0.78** | **0.83** | **0.80** |
